# COVID-19 Prevention at Institutions of Higher Education, United States, 2020 – 2021: Implementation of Nonpharmaceutical Interventions

**DOI:** 10.1101/2022.07.15.22277675

**Authors:** Sarah Moreland, Nicole Zviedrite, Faruque Ahmed, Amra Uzicanin

**Author notes:** Corresponding Author: Sarah Moreland. **Disclaimer:** The findings and conclusions in this report are those of the authors and do not necessarily represent the official position of the US Centers for Disease Control and Prevention. During the study period, SM was a fellow appointed through the Research Participation Program at the Centers for Disease Control and Prevention administered by the Oak Ridge Institute for Science and Education through an interagency agreement between the US Department of Energy and the US Centers for Disease Control and Prevention. **IRB Approval:** The project underwent ethical review at the Centers for Disease Control and Prevention’s Human Research Protections Office and was determined not to involve human subjects; it was therefore not subject to institutional review board review requirements. **Author Contributions:** Conception and design: SM; Analysis and interpretation of the data: SM, NZ, FA, AU; Drafting of the article: SM, NZ; Critical revision of the article for important intellectual content: SM, NZ, FA, AU; Final approval of the article: AU; Collection and assembly of data: SM.

## Abstract

**Background:** In early 2020, following the start of the coronavirus disease 2019 (COVID-19) pandemic, institutions of higher education (IHEs) across the United States rapidly pivoted to distance learning to reduce risk of on-campus virus transmission.

**Objective:** To explore IHE use of nonpharmaceutical interventions (NPIs) during the subsequent pandemic-affected academic year 2020–2021.

**Design:** Cross-sectional study of data collected January – June 2021. Setting: US four-year, undergraduate IHEs.

**Patients (or Participants):** All public (n=547) and a stratified random sample of private (n=300) IHEs.

**Measurements:** From IHE websites, we documented NPIs, including changes to the calendar, learning environment, housing, common areas, and dining; COVID-19 testing; and facemask protocols, and performed weighted analysis to assess congruence with the US Centers for Disease Control and Prevention (CDC) guidance for IHEs. We used weighted multivariable linear regression to explore the association between IHE characteristics and the summated number of implemented NPIs.

**Results:** Overall, 20% of IHEs implemented all surveyed CDC-recommended NPIs. The most frequently utilized were learning environment changes (91%), practiced as one or more of the following: distance or hybrid learning opportunities (98%), 6-feet spacing (60%), and reduced class sizes (51%). Additionally, 88% of IHEs specified facemask protocols, 78% physically modified common areas, and 67% offered COVID-19 testing. Among the 33% of IHEs offering ≥50% of courses in person, having <1,000 students was associated with having implemented fewer NPIs than IHEs with ≥1,000 students.

**Limitations:** Data collected from publicly available sources may introduce observation biases but allow for large sample size.

**Conclusion:** Only 1 in 5 IHEs implemented all surveyed CDC recommendations, while a majority implemented a subset. IHE size and location were associated with degree of NPI implementation. Additional research is needed to assess adherence to NPI implementation in IHE settings.

**Funding Source:** United States Centers for Disease Control and Prevention

## Introduction

In early 2020, nonpharmaceutical interventions (NPIs) were implemented across the United States (US) to slow the spread of severe acute respiratory syndrome coronavirus 2 (SARS CoV-2), the causative agent of coronavirus disease 2019 (COVID-19). These included mass gathering cancelations, travel restrictions, use of facemasks, and physical distancing, including a pivot to distance learning for both K-12 schools and institutions of higher education (IHEs) (1, 2).

By August 2020, COVID-19 incidence was highest among young adults 20-29 years of age (1). While generally at lower risk for severe disease and outcomes than older age groups, infected young adults transmit infection to others in their communities (3, 4). Simulation models suggest that the introduction of university students into a population worsened COVID-19 outcomes in the broader community between August 15, 2020, and December 31, 2020 (5). However, experiences from the US and Taiwan suggest that the safe re-opening of IHEs may be feasible with a combination of containment and mitigation strategies (6–9).

The rapid, near-universal transition to distance learning experienced by US IHEs in March 2020 was unprecedented in terms of duration and nationwide scale, thereby leaving administrators, faculty, and students without a clear path to return to on-campus operations (2). In May 2020, CDC issued guidance for IHE operations during the coming academic year, 2020–2021, that was updated regularly (10, 11).

We assessed how IHEs adapted educational instruction and other processes during the 2020– 2021 academic year and how they implemented recommended NPIs to prevent on-campus SARS-CoV-2 transmission.

## Methods

### Study sample

From the National Center for Education Statistic’s (NCES) Integrated Postsecondary Education Data System (IPEDS) (12), we identified traditional 4-year undergraduate public and private IHEs within the 50 states and DC, excluding primarily graduate, clinical, or trade programs. Data were obtained for the 2018–2019 academic year, the most recent year of data prior to the COVID-19 pandemic. All 547 public and a stratified random sample of private IHEs meeting inclusion criteria were included in the study. The universe of 1,181 private IHEs was stratified by student enrollment number as defined by IPEDS (< 1,000, 1,000 - 4,999, 5,000 - 9,999, 10,000 - 19,999, ≥ 20,000). Based on sample size calculations using a 95% confidence interval with a 5% margin of error, 26% of eligible IHEs were randomly selected within each stratum (97 of 398 IHEs, 161 of 625 IHEs, 24 of 93 IHEs, 12 of 47 IHEs, and 6 of 18 IHEs, respectively).

### Data collection

From January through June 2021, we conducted searches of publicly available online data from IHE-run websites, including those pertaining to COVID-19 response, plans to return to in-person learning, and campus announcements, to examine NPIs implemented within 547 public IHEs and the sample of 300 private IHEs in response to the ongoing COVID-19 pandemic. From IHE websites, we sought available information about NPIs implemented, with particular attention to those recommended in the October 29, 2020 update to the US Centers for Disease Control and Prevention’s (CDC) Guidance for IHEs (11). We summarized the CDC guidance into seven broad categories of interventions and designed a standardized data collection form, including fields for changes to the academic calendar, learning environment, residence halls, common spaces, and student dining; campus COVID-19 testing protocols; and facemask requirements. We collected data on NPIs specifically related to the COVID-19 pandemic rather than those routinely recommended for everyday use regardless of pandemic status, such as hand washing, cleaning, and respiratory etiquette (13). Where available, we documented how each NPI was implemented and the primary learning format utilized by the IHE. Table 1 lists all NPIs surveyed and how they were classified. Data collection for public and private IHEs was done consecutively, January 2021 – May 2021 and April 2021 – June 2021, respectively.

**Table 1.** Nonpharmaceutical interventions (NPIs) abstracted from IHE websites, derived from CDC guidance, United States, Academic Year 2020–2021.

| Category | Variable | Variable Type | Response Examples |
| --- | --- | --- | --- |
| 1. Academic Calendar |  |  |  |
|  | Has the academic calendar been changed in response to COVID-19? | Yes, No, or Not found |  |
|  | How was each campus break (Thanksgiving, winter break, spring break, etc.) changed? | Select all that apply | Shortened, lengthened, eliminated, unchanged |
| 2. Learning Environment |  |  |  |
|  | Has the learning environment been adapted in response to COVID-19? | Yes, No, or Not found |  |
|  | How has the learning environment been changed? | Select all that apply | Spacing between seats, hybrid attendance, reduced class size, other |
|  | In what format(s) are classes being offered? | Select all that apply | In person, hybrid, distance learning |
| 3. On-Campus Housing |  |  |  |
|  | Has the capacity of residence halls been changed or reduced? | Yes, No, Not found, or Does not apply |  |
|  | How has residence hall capacity been changed? | Select all that apply | Single-occupancy rooms, reduced capacity rooms, designated isolation spaces, other |
|  | Have residence hall guest and visitor policies been changed? | Yes, No, Not found, or Does not apply |  |
|  | How have residence hall guest policies been changed? | Select all that apply | Eliminate visitation, limit number of visitors, restrict visitors based on residence, other |
| 4. Campus Common Spaces |  |  |  |
|  | Have common spaces been physically changed or restricted? | Yes, No, or Not found |  |
|  | How have common spaces been changed? | Select all that apply | Removal of furniture, signage and floor stickers, closure of spaces, limit capacity, other |
| 5. Campus Dining Services |  |  |  |
|  | Have dining halls and services been changed or limited? | Yes, No, or Not found |  |
|  | How have dining services been changed? | Select all that apply | Limit capacity, to-go dining, installation of partitions, removal of self-serve options, other |
| 6. COVID-19 Testing |  |  |  |
|  | Does the IHE offer COVID-19 testing on campus? | Yes, No, or Not found |  |
|  | Who is eligible to utilize campus COVID-19 testing services? | Select all that apply | Residential students, all students, staff, non-campus affiliates |
|  | What criteria must be met to receive a COVID-19 test? | Select all that apply | Asymptomatic, suspected contact, confirmed contact, symptomatic, no criteria |
|  | When, if ever, is COVID-19 testing required? | Select all that apply | Surveillance testing, upon arrival for the semester |
|  | How did COVID-19 testing availability change between semesters? | More available, Less available, No change |  |
| 7. Facemask Requirements |  |  |  |
|  | Where are facemasks required on campus? | Select all that apply | Indoor, outdoor (where 6ft cannot be maintained) |
|  | Who do facemask requirements apply to? | Select all that apply | Students, staff, visitors |

\* NPIs derived from October 29, 2020 update of US Centers for Disease Control and Prevention Guidance for IHEs.

### Final dataset

From the NCES IPEDS dataset, we collected data on the characteristics of IHEs included in our study, including affiliation (public vs. private), student enrollment, degree of urbanization, and location information (county name and code). IHEs were categorized into four geographic regions as defined by the US Census Bureau (14). IHEs were further categorized by size into small (>1,000 students), medium (1,000 – 9,999 students), and large (≥10,000 students). We obtained publicly available county-level data on COVID-19 cases from January 2020 – June 2021 from the COVID-19 Data Repository by the Center for Systems Science and Engineering (CSSE) at Johns Hopkins University (15, 16). The national incidence rate per 100,000 population (CI) for each week during the period May 2020 – June 2021 was calculated using the summated reported daily COVID-19 case counts and the US Census Bureau 2020 estimated residential population (15–17). To visualize national COVID-19 trends in the context of the 2020–2021 academic year, IHE academic calendar dates were collected and summarized by median and IQR dates for semester start and end, and each break within the first semester, classified as August through December 2020, and the second semester, classified as January through May 2021, and illustrated graphically along with US weekly COVID-19 incidence (Figure 1). To simplify variation in calendar structure between IHEs, our analysis was restricted to dates encompassing the common two-semester academic year, excluding summer or other abbreviated terms.

**Figure 1.**
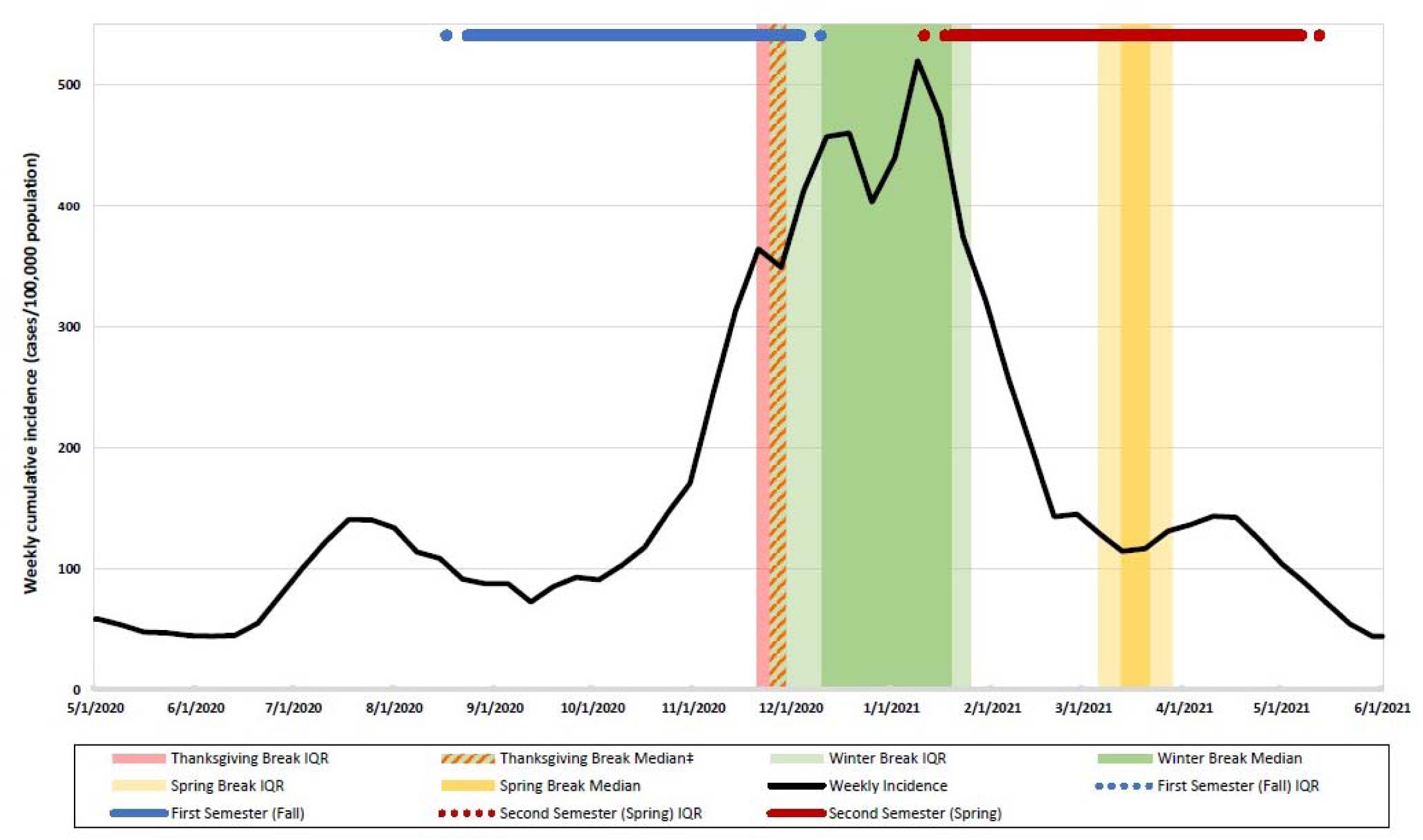
Timeline of institution of higher education (IHE) academic calendars* and weekly national COVID-19 incidence†, United States, Academic Year 2020–2021. *2020-2021 Academic calendar dates collected from each IHE website. Median (50 percentile) and IQR (25 and 75 percentile) were calculated for milestone dates. †Weekly Incidence (cases per week per 100,000 people) derived from JHU CSSE COVID-19 Data and U.S. Census Bureau 2020 population estimation. ‡ Thanksgiving median and IQR, and winter break IQR dates encompass November 25-29, 2020.

### Analysis and visualization

Descriptive statistics were calculated to summarize characteristics of the sampled IHEs, including affiliation, enrollment, US Census Bureau Region, and urbanicity. The number of NPIs implemented was summated to create a score from 0-7, representing a level of compliance with the seven categories of NPIs shown in Table 1 (11). For each NPI category, a score of 1 was assigned if it was implemented through at least one modality, such as the examples listed in Table 1, and a score of 0 was assigned if it was not implemented or if implementation was unknown. To generalize findings to all US IHEs, we performed weighted analysis to assess congruence with the guidance. Sampling weights representing the inverse of the sampling fractions were used to account for the unequal probability of selection into the sample (18). For IHEs that offered ≥50% courses in-person, we used weighted multivariable linear regression to explore the associations between IHE characteristics and the summated number of implemented NPIs. Weighted multivariable logistic regression was performed to explore associations between IHE characteristics and individual NPIs among IHEs offering ≥50% courses in person.

Analysis was conducted using SAS 9.4. Figures were created in Tableau, Microsoft Power BI, and Microsoft Excel.

## Results

### Characteristics of the study sample

A total of 847 IHEs were sampled from the universe of 1,728 IHEs, including all 547 (32%) public and 300 of the 1,181 (68%) private institutions (Table 2). Of the four US Census Bureau regions, the South contained the highest number of IHEs, and the West contained the fewest. Half of the sampled IHEs were in cities, and the fewest were in rural areas. IHEs were categorized based on student enrollment: 24% enrolled under 1,000 students, 44% enrolled 1,000 – 4,999 students, 13% enrolled 5,000 – 9,999 students, 10% enrolled 10,000 – 19,999 students, and 9% enrolled 20,000 or more students.

**Table 2.** Characteristics of sampled IHEs by institution size*, United States, Academic Year 2020–2021 (N=847)

| Institution size category* | Total | Under 1,000 students | 1,000 - 4,999 students | 5,000 - 9,999 students | 10,000 - 19,999 students | 20,000 students and above | p-value |
| --- | --- | --- | --- | --- | --- | --- | --- |
| Number of IHEs, n (%)† | 847 (100) | 118 (24) | 302 (44) | 152 (13) | 139 (10) | 136 (9) |  |
| Affiliation, n (%)† |  |  |  |  |  |  | <0.0001 |
| Public | 547 (32) | 21 (5) | 141 (18) | 128 (58) | 127 (73) | 130 (88) |  |
| Private | 300 (68) | 97 (95) | 161 (82) | 24 (42) | 12 (27) | 6 (12) |  |
| United States Census Region, n (%)† |  |  |  |  |  |  | <0.0001 |
| Northeast | 204 (26) | 38 (25) | 79 (28) | 45 (35) | 29 (23) | 13 (13) |  |
| Midwest | 213 (28) | 33 (32) | 80 (30) | 35 (24) | 37 (26) | 28 (19) |  |
| South | 305 (34) | 35 (33) | 112 (34) | 53 (29) | 50 (35) | 55 (39) |  |
| West | 125 (12) | 12 (11) | 31 (8) | 19 (13) | 23 (15) | 40 (30) |  |
| Urbanicity‡, n (%)† |  |  |  |  |  |  | <0.0001 |
| Rural | 50 (7) | 18 (14) | 29 (8) | 3 (1) | 0 (0) | 0 (0) |  |
| Town | 186 (18) | 18 (15) | 89 (22) | 42 (20) | 29 (17) | 8 (5) |  |
| Suburban | 194 (25) | 33 (25) | 69 (26) | 49 (31) | 29 (20) | 23 (16) |  |
| City | 417 (50) | 49 (45) | 115 (44) | 67 (47) | 81 (63) | 105 (79) |  |

†The universe of 1,181 private IHEs was stratified by institution size category. Based on sample size calculations using a 95% confidence interval with a 5% margin of error, 26% of eligible IHEs were randomly selected within each stratum (<1,000: 97 of 398 IHEs, 1,000 – 4,999: 161 of 625 IHEs, 5,000 – 9,999: 24 of 93 IHEs, 10,000 – 19,999: 12 of 47 IHEs, and ≥20,000: 6 of 18 IHEs). Weights represent the inverse of the sampling fractions. n (%) represents the unweighted number (n) and the weighted percent of the universe of 1,728 IHEs.
‡Level of urbanization based on urban boundaries by Census GEO as reported by the Integrated Postsecondary Education Data System.

### CDC Guidance compliance

The October 29, 2020, CDC Guidance for IHEs was comprised of 7 broad interventions (11), and their implementation resulted in at least 24 distinct modalities in practice, summarized in Appendix Table 1. All recommended NPIs were addressed and implemented through at least one modality by 20% of IHEs, and 4% implemented none (Appendix Table 1). IHEs implementing all 7 NPIs more often had higher enrollment, while those implementing none most often had <1,000 students. The four-NPI combination of changes to the learning environment, common spaces, dining, and facemask protocols were implemented by 62% of IHEs. Figure 2 illustrates the implementation of NPIs by institution size category.

**Figure 2.**
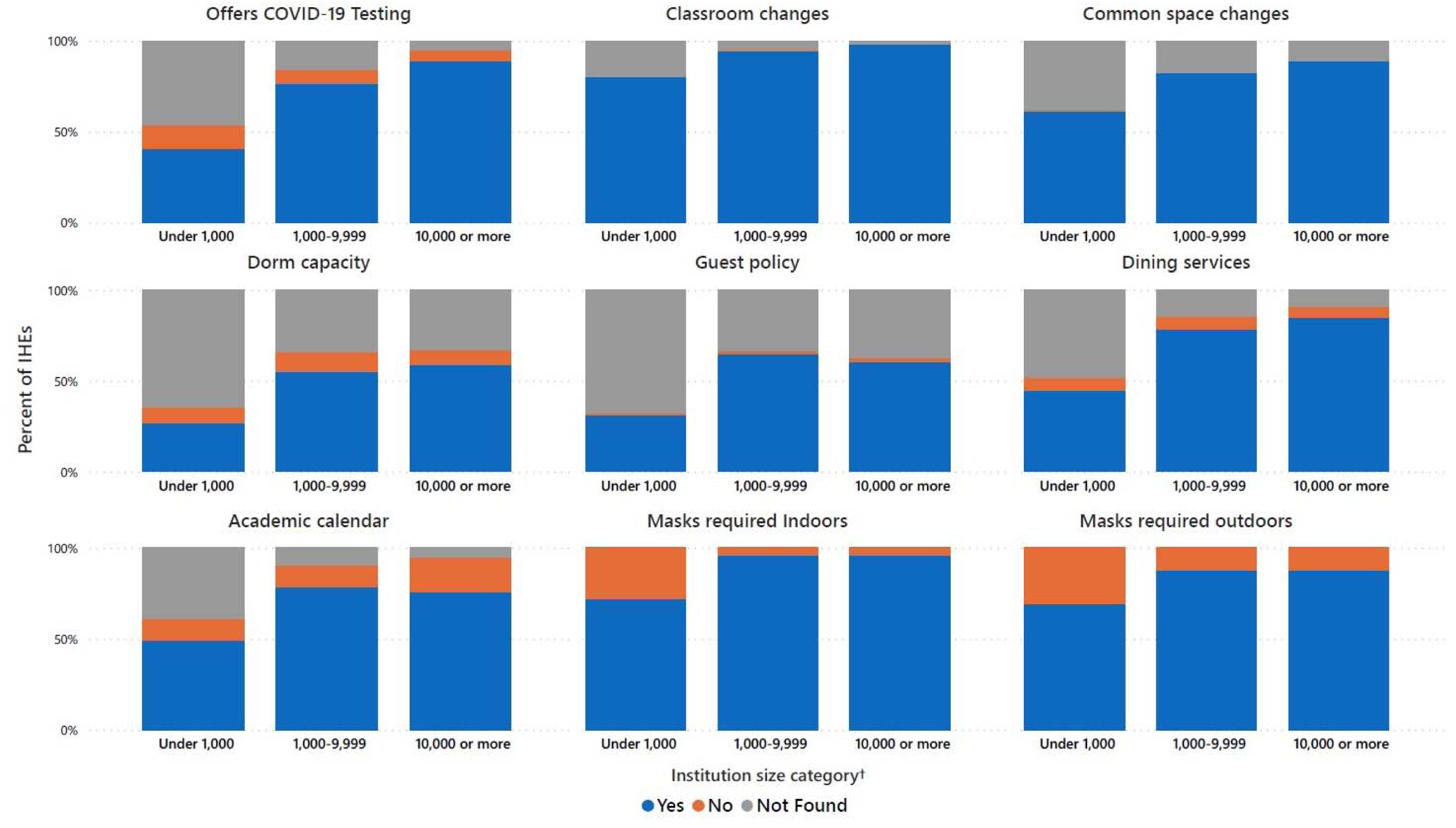
Nonpharmaceutical interventions (NPIs) implemented* by institutions of higher education (IHEs) (N = 847) in response to the COVID-19 pandemic by enrollment size category†, United States, Academic Year 2020–2021. * NPIs derived from CDC Guidance October 29, 2020 update of US Centers for Disease Control and Prevention Guidance for IHEs † Institution size category based on total students enrolled for credit as reported by the Integrated Postsecondary Education Data System.

### Primary learning method

The primary format of course delivery was identified for 393 IHEs, of which 38% offered courses <50% in person, 26% offered courses >50% in person, and 36% offered courses roughly 50% through distance learning and 50% in person (Appendix Table 1). IHEs that offered ≥50% of courses in person were most often smaller by student enrollment and in the Midwest (Figure 3, Appendix Figure 1). In multivariable regression, IHEs having 1,000-4,999 students enrolled versus <1,000, being in the Midwest versus the Northeast, and being in a town versus urban were more likely to offer ≥50% of courses in person (Appendix Table 2). IHEs in the West versus Northeast were less likely to have offered ≥50% of courses in person.

**Figure 3.**
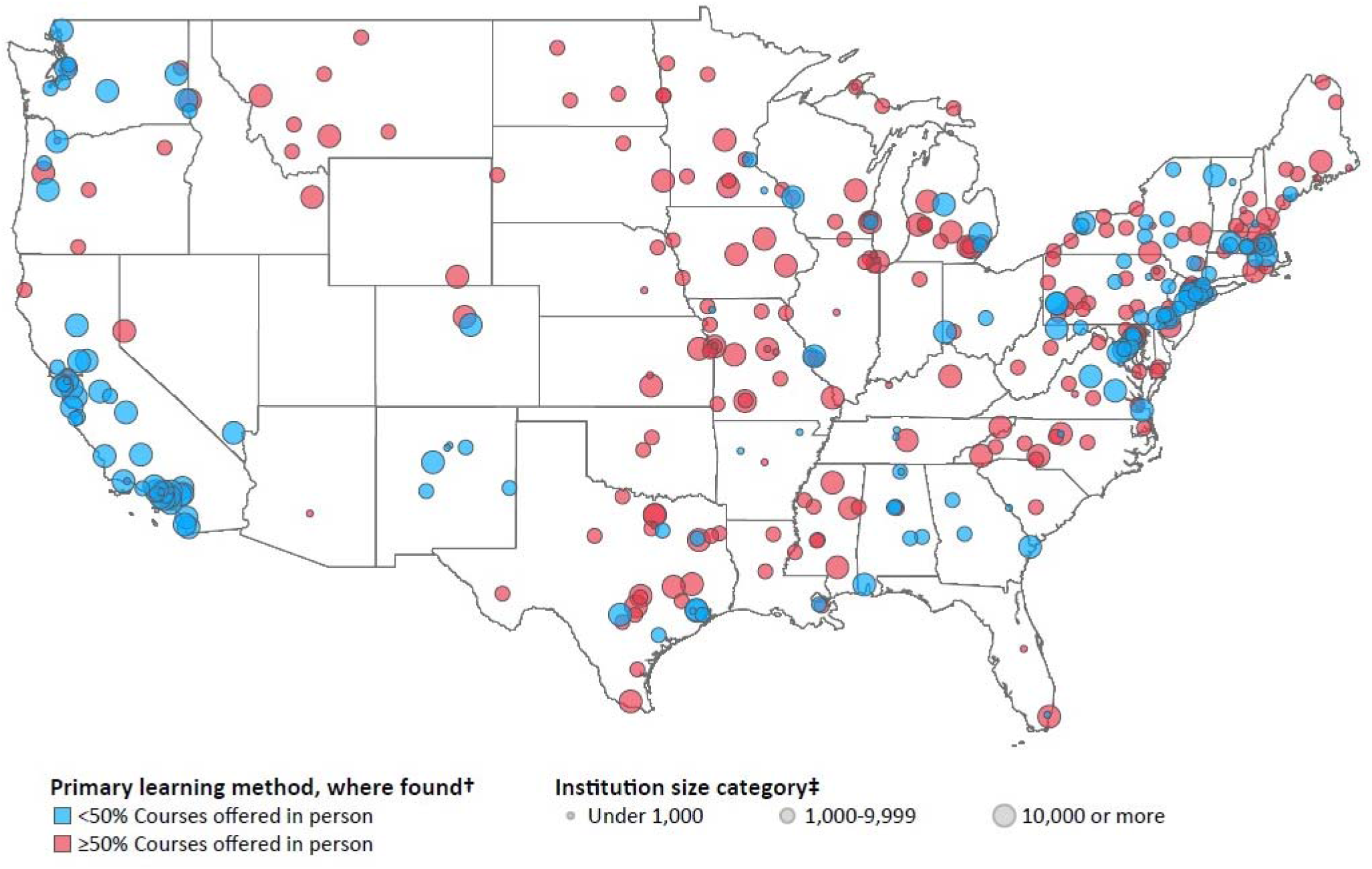
Geographical distribution* of primary learning method† offered by institutions of higher education (IHEs) (N=393), by institutional size category‡, United States, Academic Year 2020–2021. * Geographical distribution of primary learning method with size of the point representing institution enrollment size category. Generated with Tableau Software † 227 IHEs offer courses ≥50% in person. 166 IHEs offer courses <50% in person. Primary learning method not found for 454 IHEs. ‡ Institution size category based on total students enrolled for credit as reported by the Integrated Postsecondary Education Data System.

Among the subset of IHEs that reported offering ≥50% courses in person, the number of NPIs implemented was higher among IHEs in the Northeast compared to the other regions and among IHEs having 1,000-4,999 and 10,000-19,999 students than those with <1,000 students (Table 3). When examined visually by affiliation, private IHEs exhibited wider variation in the number of NPIs implemented than public IHEs, most notably in IHEs with under 1,000 students (Appendix Figure 2).

**Table 3.**
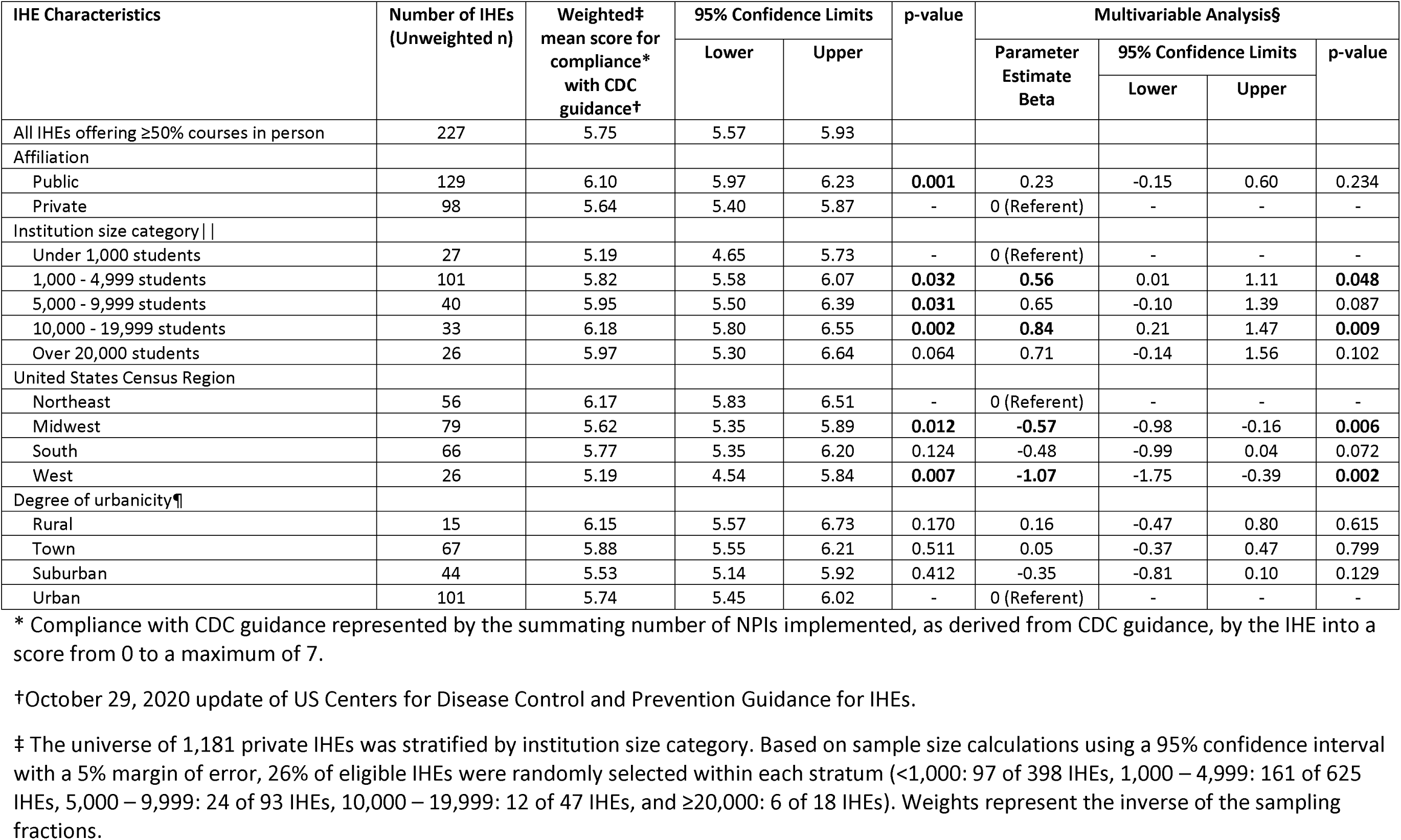
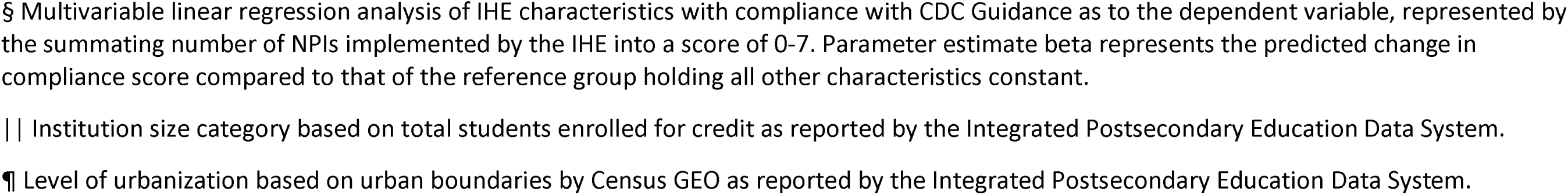
IHE characteristics associated with compliance* with CDC Guidance† among IHEs offering primarily in-person learning (N=227), United States, Academic Year 2020–2021.

| IHE Characteristics | Number of IHEs<br>(Unweighted n) | Weighted‡<br>mean score for<br>compliance*<br>with CDC<br>guidance† | 95% Confidence Limits |  | p-value | Multivariable Analysis§ |  |  |  |
| --- | --- | --- | --- | --- | --- | --- | --- | --- | --- |
|  |  |  | Lower | Upper |  | Parameter<br>Estimate<br>Beta | 95% Confidence Limits |  | p-value |
|  |  |  |  |  |  |  | Lower | Upper |  |
| All IHEs offering ≥50% courses in person | 227 | 5.75 | 5.57 | 5.93 |  |  |  |  |  |
| Affiliation |  |  |  |  |  |  |  |  |  |
| Public | 129 | 6.10 | 5.97 | 6.23 | 0.001 | 0.23 | -0.15 | 0.60 | 0.234 |
| Private | 98 | 5.64 | 5.40 | 5.87 | - | 0 (Referent) | - | - | - |
| Institution size category \|\| |  |  |  |  |  |  |  |  |  |
| Under 1,000 students | 27 | 5.19 | 4.65 | 5.73 | - | 0 (Referent) | - | - | - |
| 1,000 - 4,999 students | 101 | 5.82 | 5.58 | 6.07 | 0.032 | 0.56 | 0.01 | 1.11 | 0.048 |
| 5,000 - 9,999 students | 40 | 5.95 | 5.50 | 6.39 | 0.031 | 0.65 | -0.10 | 1.39 | 0.087 |
| 10,000 - 19,999 students | 33 | 6.18 | 5.80 | 6.55 | 0.002 | 0.84 | 0.21 | 1.47 | 0.009 |
| Over 20,000 students | 26 | 5.97 | 5.30 | 6.64 | 0.064 | 0.71 | -0.14 | 1.56 | 0.102 |
| United States Census Region |  |  |  |  |  |  |  |  |  |
| Northeast | 56 | 6.17 | 5.83 | 6.51 | - | 0 (Referent) | - | - | - |
| Midwest | 79 | 5.62 | 5.35 | 5.89 | 0.012 | -0.57 | -0.98 | -0.16 | 0.006 |
| South | 66 | 5.77 | 5.35 | 6.20 | 0.124 | -0.48 | -0.99 | 0.04 | 0.072 |
| West | 26 | 5.19 | 4.54 | 5.84 | 0.007 | -1.07 | -1.75 | -0.39 | 0.002 |
| Degree of urbanicity¶\|\| |  |  |  |  |  |  |  |  |  |
| Rural | 15 | 6.15 | 5.57 | 6.73 | 0.170 | 0.16 | -0.47 | 0.80 | 0.615 |
| Town | 67 | 5.88 | 5.55 | 6.21 | 0.511 | 0.05 | -0.37 | 0.47 | 0.799 |
| Suburban | 44 | 5.53 | 5.14 | 5.92 | 0.412 | -0.35 | -0.81 | 0.10 | 0.129 |
| Urban | 101 | 5.74 | 5.45 | 6.02 | - | 0 (Referent) | - | - | - |

\* Compliance with CDC guidance represented by the summing number of NPIs implemented, as derived from CDC guidance, by the IHE into a score from 0 to a maximum of 7.
†October 29, 2020 update of US Centers for Disease Control and Prevention Guidance for IHEs.
‡ The universe of 1,181 private IHEs was stratified by institution size category. Based on sample size calculations using a 95% confidence interval with a 5% margin of error, 26% of eligible IHEs were randomly selected within each stratum (<1,000: 97 of 398 IHEs, 1,000 – 4,999: 161 of 625 IHEs, 5,000 – 9,999: 24 of 93 IHEs, 10,000 – 19,999: 12 of 47 IHEs, and ≥20,000: 6 of 18 IHEs). Weights represent the inverse of the sampling fractions.
§ Multivariable linear regression analysis of IHE characteristics with compliance with CDC Guidance as to the dependent variable, represented by the summing number of NPIs implemented by the IHE into a score of 0-7. Parameter estimate beta represents the predicted change in compliance score compared to that of the reference group holding all other characteristics constant.
|| Institution size category based on total students enrolled for credit as reported by the Integrated Postsecondary Education Data System.
¶ Level of urbanization based on urban boundaries by Census GEO as reported by the Integrated Postsecondary Education Data System.

### Academic Calendar

Over two-thirds of IHEs made changes to their academic calendars in response to COVID-19 (Appendix Table 1). Figure 1 summarizes common IHE academic breaks. Among the 418 IHEs with available information, 77% lengthened Thanksgiving break, nearly all of which extended it into winter break and did not return to in-person instruction for the remainder of the calendar year. Where identified (386 IHEs), half of IHEs specifically lengthened winter break with additional days off, a period of distance learning, or a combination of the two. Among the 511 IHEs where information was identified, 68% eliminated spring break.

### Learning Environment

The most frequently utilized NPI, implemented by 91% of IHEs, was changes to the learning environment which was practiced most often among this subset as one or more of the following: distance or hybrid learning options (98%), 6-feet spacing (60%), reduced class sizes (51%), and alternating classroom attendance (38%) (Appendix Table 1). Only one IHE explicitly did not implement any changes, and information was not identified for 9%.

### Housing

Residential housing capacity policies were changed in 50% of IHEs, not changed in 11%, and not identified in 36% (Appendix Table 1). On-campus housing was not offered in 3%. Overall housing capacity or density was reduced in 55% of IHEs, and, notably, dorms were limited to single occupancy in 20%. Residential housing guest policies were changed in 58% of IHEs, not changed in 2%, and not identified in 37% (Appendix Table 1).

### Common Spaces

Changes to common campus spaces were made by 78% of IHEs, practiced most often among this subset as one or more of the following: reduced capacity (74%), physical guides (58%), such as signs or floor markers, or physical modifications (54%), such as furniture removal or use of partitions (Appendix Table 1). Changes were most often made by IHEs with ≥1,000 students. Only one IHE explicitly did not implement any changes to common spaces, and information was not identified for 22% IHEs.

### Dining Services

Dining services were modified by 72% of IHEs, among which most implemented one or more of the following methods: increased to-go meal options (73%), reduced indoor seating capacity (67%), and physical modifications, such as furniture removal or use of partitions (45%) or elimination of self-service options (48%) (Appendix Table 1). IHEs with ≥1,000 students made changes to dining most often. Dining services were explicitly unchanged in 3% of IHEs, and information was not identified for 25%.

### COVID-19 Testing

Over two-thirds of IHEs were found to offer on-campus COVID-19 testing, most often larger IHEs (Appendix Table 1). Among IHEs that offered on-campus COVID-19 testing, testing was required by 83% under one or more circumstances, such as upon arrival for the semester or ongoing surveillance testing. Among IHEs learning ≥50% in person, those with testing requirements were significantly more likely to have over 10,000 students compared to those with under 1,000 students (Appendix Table 3). Testing requirements were more common in the Northeast than in the Midwest, and in urban than suburban areas.

### Facemask Policies

Facemasks were required by 88% of IHEs when indoors on campus property (Appendix Table 1). Similarly, 82% of IHEs required facemasks outdoors whenever a 6-foot distance could not be maintained. Policies requiring facemasks were least prevalent both indoors and outdoors in IHEs with <1,000 students.

## Discussion

### Main Findings

While 96% of sampled IHEs deployed NPIs during the 2020–2021 academic year in response to the ongoing COVID-19 pandemic, only 1 in 5 (20%) comprehensively complied with the CDC guidance. Degree of compliance was associated with both IHE size and location, with larger IHEs (≥1,000 students) and those located in the Northeast (Connecticut, Maine, Massachusetts, New Hampshire, New Jersey, New York, Pennsylvania, Rhode Island, and Vermont) being most likely to have high compliance. Conversely, the lowest compliance was most often observed in private IHEs and those located in the West (Arizona, Alaska, California, Colorado, Hawaii, Idaho, Nevada, New Mexico, Oregon, Washington, and Wyoming).

IHEs typically harbor high attack rates during outbreaks of respiratory illness; however, little comprehensive data on IHE outbreak response exist, including during the COVID-19 pandemic (19, 20). Although measures introduced at the onset of the COVID-19 pandemic, such as distance learning and physical distancing, continue to be used (2, 21, 22), the scientific evidence base on their effects and effectiveness in reducing respiratory virus transmission (2022) is still evolving, and it was even more limited in early 2020 when the initial CDC IHE guidance was issued (11). IHEs that re-opened for the 2020 – 2021 academic year reported implementation of a more streamlined set of NPIs across the US and globally, which included distancing in the learning environment, use of facemasks, and COVID-19 testing protocols (6, 8, 9, 23–28). However, much of the currently available literature is focused on the implementation of few or individual NPIs rather than a comprehensive evaluation of layered NPIs in COVID-19 prevention (25, 27–29). However, although most currently available studies are limited to specific populations and not fully generalizable to IHEs across the US (6, 8, 9, 23), they suggest that regardless of size or location, IHEs must take a multifaceted approach and timely implement combinations of appropriate NPIs to effectively reduce transmission of SARS-CoV-2 on campus (9, 30, 31).

Among IHEs where the primary learning method was identified, about one-third reported offering classes primarily through distance education. Our data suggest that large or public IHEs were more likely to have primarily offered distance learning, while also being more likely to have made physical changes to the learning environment when utilizing in-person instruction. Modalities of learning environment changes such as distanced seating configurations, hybrid schedules, and reduced class sizes are widely used to maximize in-person opportunities while minimizing the risk of SARS-CoV-2 transmission by decreasing student contact (30–34). Notably, distance learning has been reported to create significant challenges for students, particularly those of low- and middle-income backgrounds or those who rely on on-campus facilities and services for accessible technology, quiet space, or stable housing (35–38). Reasons for preference in learning method are complex and are beyond the scope of this research; however, factors such as logistic capabilities, funding, local politics, and perceptions of distance learning may have played roles in decision making (2, 39, 40). Therefore, IHEs should carefully consider the benefits of in-person operations, their ability to implement NPIs to facilitate safe in-person operations, and the health of IHE populations and risks for surrounding communities during a severe pandemic such as COVID-19 (28, 41).

Two-thirds of IHEs made changes to the academic calendar in response to COVID-19. Lengthening of breaks, either by transitioning to a period of distance learning or by adjusting academic start or end dates, acts as a temporary IHE dismissal allowing for longer periods between mixing of campus populations with outside populations and re-congregation on campus. These have been employed both proactively and reactively by IHEs to reduce campus-based SARS-CoV-2 transmission (6, 8, 23). Elimination of breaks – a measure that is not a part of CDC pre-pandemic guidelines or pandemic guidance – reduces the potential number of consecutive student days away from campus in the middle of the term, theoretically reducing opportunities to bring new viral lineages onto IHE campuses and surrounding communities (29). For example, a large cluster of COVID-19 cases on a Chicago university campus following spring break revealed roughly two-thirds of sequenced cases originated outside of Chicago (42). It is unlikely, however, that the break elimination would reduce already established virus transmission.

Despite widespread efforts to reduce density and limit visitation as IHEs re-opened campus for the 2020–2021 academic year, many reported COVID-19 outbreaks linked to residential housing due to the difficulty of maintaining and enforcing physical distancing measures in those spaces (8, 23, 27, 43, 44). IHE policies restricting congregation in campus spaces and housing may be difficult to enforce and do not extend to off-campus student gatherings.

Our data suggest that larger IHEs were more likely than smaller IHEs to require COVID-19 testing. On-campus surveillance testing tends to be the most expensive and logistically demanding COVID-19 mitigation measure, and frequent deployment may pose a challenge, particularly to small IHEs, which receive proportionally less enrollment-based funding or may lack extensive laboratory infrastructure (26, 39, 45). Because testing is an important public health tool to reduce SARS-CoV2 transmission in dense congregate settings such as IHEs, reasons for limited compliance with testing recommendations should be elucidated and addressed (27, 46, 47).

Compared to laboratory testing, facemasks pose less of a financial and logistical burden on IHEs (40). A majority of IHEs required facemasks indoors and outdoors wherever six-foot spacing could not be maintained. The encouraged or required use of facemasks has been consistently cited as a core NPI utilized by IHEs in response to COVID-19 (23, 24, 48, 49). Observational reports reveal high levels of compliance with facemask mandates on IHE campuses (25), and they remain an important measure for reducing SARS-CoV2 transmission, especially in dense indoor settings.

### Limitations

Our results should be considered in the context of at least four limitations. First, publicly available data are not guaranteed to fully reflect all NPIs implemented within IHEs. However, our method allowed us to obtain the data on recommended NPIs quickly from a wide variety of measures from a large sample of public and private institutions. Additionally, the CDC Guidance for IHEs made broad recommendations intended to be applied by a wide variety of IHEs; as such, it left room for interpretation in how IHEs implemented the guidance, which in turn posed a challenge in the data collection process. If an NPI was not explicitly mentioned on an IHE website, it was recorded as “not found,” minimizing assumptions and misclassification biases. Public IHEs were more likely to be larger and have publicly available COVID-19 information than private IHEs, which may contribute to the size- or affiliation-related variations in documented NPIs.

Second, a team of several people was responsible for data collection, which can lead to systematic bias. Response frequencies for each NPI were compared within and between each member of the data collection team to ensure patterns or outliers were consistent with the original data sources. Acceptable patterns were found to be due to university systems or state and local governments issuing streamlined guidance for all IHEs under their influence.

Third, data were collected during the second semester of the 2020–2021 academic year, meaning changes made during the first semester may have been missed. Although this has the potential to introduce response bias as publicly available information evolves over time, data were collected systematically, ensuring the data described measures representing NPIs actively implemented or how they evolved throughout the entire 2020–2021 academic year.

Finally, this study aims to evaluate and describe the steps taken by IHEs to minimize SARS-CoV-2 transmission on campus and does not account for adherence to the documented NPIs, or off-campus behavior.

## Conclusions

In conclusion, the COVID-19 pandemic had an unprecedented impact on IHEs across the US, where there has previously been little comprehensive data on respiratory illness outbreaks and responses. Some NPIs were widely implemented as a means of reducing SARS-CoV2 transmission within IHEs, but the degree of compliance with recommended NPIs varied by IHE size and location. Further research is needed to understand the reasons for suboptimal compliance, including financial and logistical challenges, and to address barriers to the implementation of recommended NPIs. In future studies evaluating the effectiveness of these measures in IHE settings, level of compliance with NPIs as described here, as well as levels of adherence and the impact of off-campus behaviors should be taken into consideration. As IHEs continue to navigate the ongoing COVID-19 pandemic, they must adapt their normal operations to prioritize the health of students and staff through layered COVID-19 prevention, including vaccination, timely case detection through testing and tracing, and continued use of NPIs as feasible and appropriate for the local epidemiologic situation. In addition to the discussed NPIs, for sustainable control of respiratory infections, including COVID-19, IHEs are encouraged to improve campus ventilation infrastructure, increase opportunities for physical distancing such as open-air study spaces, exercise flexibility in distance learning and staying home when sick, and promote the consistent use of facemasks during the seasonal waves of respiratory infections, particularly for anyone (students, staff, visitors) who is experiencing respiratory symptoms (10).

## Data Availability

All data produced in the present study are available upon reasonable request to the authors.

## Appendix

**Appendix Figure 1.**
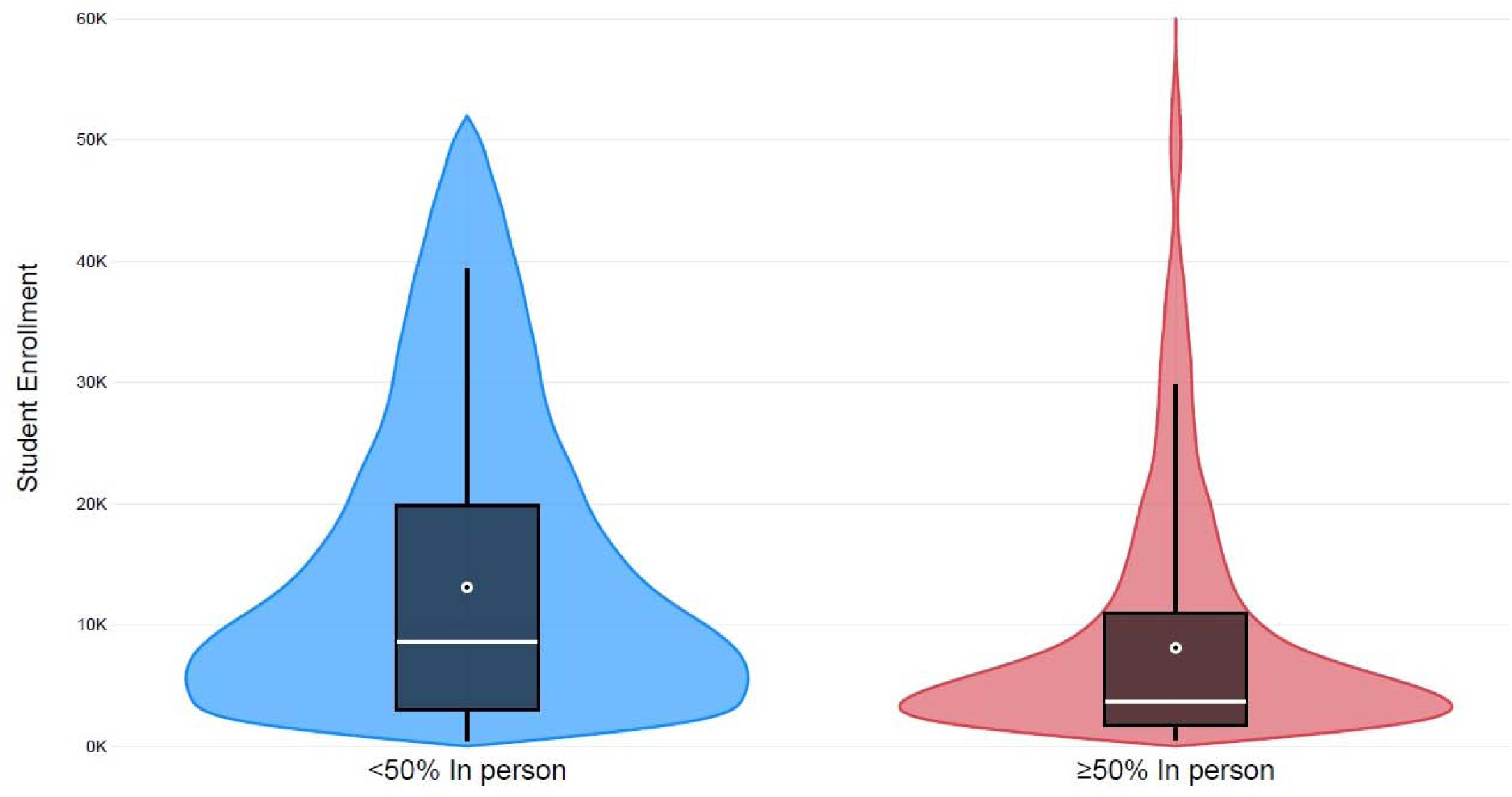
Distribution* of institution of higher education (IHE) student enrollment by primary course format† offered by IHEs (N=393), United States, Academic Year 2020 – 2021. * Violin plot of student enrollment kernel density distribution within primary learning formats. Boxplot dot and line represent the mean and median enrollment, respectively. Generated in Microsoft Power BI. † 227 IHEs offer courses ≥50% in person. 166 IHEs offer courses <50% in person. Primary learning method not found for 454 IHEs.

**Appendix Table 1.** NPIs implemented by IHEs (N=847) as derived from CDC Guidance in response to the COVID-19 pandemic by institution size category†, United States, Academic Year 2020–2021.

| Institution size category† | Total | Under 1,000 students | 1,000 - 4,999 students | 5,000 - 9,999 students | 10,000 - 19,999 students | 20,000 students and above |
| --- | --- | --- | --- | --- | --- | --- |
| Number of IHEs, n (%)‡ | 847 (100) | 118 (24) | 302 (44) | 152 (13) | 139 (10) | 136 (9) |
| Selected combinations of NPIs implemented by IHEs, n (%)‡ |  |  |  |  |  |  |
| 7 Nonpharmaceutical interventions |  |  |  |  |  |  |
| All surveyed CDC guidelines* | 182 (20) | 9 (7) | 70 (24) | 38 (25) | 27 (24) | 38 (28) |
| 5 Nonpharmaceutical interventions |  |  |  |  |  |  |
| COVID-19 testing, masks, changes to classroom, common space, and dining | 465 (49) | 27 (21) | 160 (55) | 89 (56) | 92 (68) | 97 (70) |
| Masks, changes to academic calendar, classroom, common space, and dining | 433 (47) | 30 (25) | 159 (52) | 84 (56) | 82 (62) | 78 (55) |
| 4 Nonpharmaceutical interventions |  |  |  |  |  |  |
| Masks, changes to classroom, common space, and dining | 557 (62) | 41 (35) | 204 (70) | 104 (67) | 108 (79) | 100 (72) |
| 0 Nonpharmaceutical interventions | 19 (4) | 15 (14) | 4 (2) | 0 (0) | 0 (0) | 0 (0) |
| Primary format of course delivery, n (%)‡ |  |  |  |  |  |  |
| Primary format identified | 393 (52) | 48 (53) | 138 (52) | 72 (53) | 68 (56) | 67 (48) |
| Mostly distance learning (<50% in person) | 166 (38) | 21 (42) | 37 (27) | 32 (39) | 35 (53) | 41 (58) |
| Balanced (50% in person and 50% distance learning) | 153 (36) | 16 (32) | 58 (37) | 32 (44) | 27 (38) | 20 (31) |
| Mostly in person learning (>50% in person) | 74 (26) | 11 (26) | 43 (36) | 8 (17) | 6 (9) | 6 (11) |
| Changes in class delivery between academic terms |  |  |  |  |  |  |
| More distance learning | 27 (5) | 5 (6) | 8 (4) | 5 (4) | 3 (3) | 6 (8) |
| No change | 164 (39) | 20 (43) | 51 (36) | 34 (46) | 29 (15) | 30 (42) |
| More in person learning | 55 (17) | 7 (16) | 20 (18) | 11 (20) | 8 (15) | 9 (15) |
| Not found | 147 (39) | 16 (34) | 59 (43) | 22 (30) | 28 (48) | 22 (34) |
| Primary format not identified | 401 (48) | 47 (47) | 144 (48) | 75 (47) | 67 (44) | 68 (52) |
| 1. Made changes to Academic Calendar, n (%)‡ |  |  |  |  |  |  |
| Yes | 623 (67) | 58 (42) | 236 (73) | 120 (78) | 101 (76) | 108 (77) |
| Specific changes to academic breaks (where changes are not unknown) |  |  |  |  |  |  |
| Lengthening of Thanksgiving Break (n=418) | 323 (77) | 42 (87) | 119 (78) | 64 (72) | 43 (75) | 54 (70) |
| Does not return to in-person learning after Thanksgiving break | 315 (76) | 42 (82) | 116 (76) | 63 (72) | 41 (75) | 53 (68) |
| Lengthening of Winter Break (n=386) | 200 (51) | 27 (53) | 65 (46) | 43 (60) | 33 (51) | 32 (56) |
| Elimination of Spring Break (n=511) | 341 (69) | 35 (71) | 131 (70) | 75 (77) | 49 (66) | 51 (58) |
| No | 120 (14) | 14 (14) | 32 (12) | 23 (16) | 29 (17) | 22 (16) |
| Unknown | 104 (19) | 46 (44) | 34 (15) | 9 (6) | 9 (7) | 6 (7) |
| 2. Made changes to learning environment, n (%)‡ |  |  |  |  |  |  |
| Yes | 794 (91) | 95 (78) | 282 (93) | 147 (96) | 135 (96) | 135 (99) |
| Offered distance or hybrid learning | 783 (98) | 91 (95) | 278 (98) | 144 (96) | 135 (100) | 135 (100) |
| Physical changes to the classroom |  |  |  |  |  |  |
| Space seating 6ft apart | 465 (60) | 43 (48) | 177 (66) | 92 (65) | 81 (57) | 72 (54) |
| Reduced class size | 413 (51) | 39 (41) | 150 (53) | 83 (54) | 74 (57) | 67 (50) |
| Alternating attendance of students in classes | 297 (38) | 27 (28) | 119 (43) | 48 (32) | 59 (49) | 44 (33) |
| No | 1 (0) | 0 (0) | 1 (0) | 0 (0) | 0 (0) | 0 (0) |
| Unknown | 52 (9) | 23 (22) | 19 (7) | 5 (4) | 4 (4) | 1 (1) |
| 3. Made changes to Student Housing, n (%)‡ |  |  |  |  |  |  |
| Residential housing capacity |  |  |  |  |  |  |
| Yes | 445 (50) | 32 (28) | 157 (54) | 94 (59) | 82 (62) | 80 (58) |
| Reduced housing density and/or capacity | 228 (55) | 14 (44) | 83 (57) | 47 (55) | 42 (58) | 42 (53) |
| Single occupancy rooms | 108 (20) | 6 (19) | 34 (17) | 22 (19) | 22 (26) | 24 (30) |
| No | 82 (11) | 10 (10) | 35 (12) | 14 (13) | 11 (8) | 12 (9) |
| Unknown | 301 (36) | 69 (57) | 105 (32) | 42 (27) | 42 (27) | 43 (32) |
| IHE does not offer student housing | 19 (3) | 7 (5) | 5 (2) | 2 (1) | 4 (3) | 1 (1) |
| Guest policy |  |  |  |  |  |  |
| Yes | 497 (58) | 37 (32) | 198 (69) | 96 (66) | 80 (61) | 86 (64) |
| Did not allow guests in living areas | 257 (50) | 16 (45) | 99 (49) | 50 (50) | 41 (50) | 51 (61) |
| Limited number of guests | 120 (22) | 7 (19) | 47 (22) | 25 (30) | 19 (18) | 22 (26) |
| No | 14 (2) | 1 (1) | 6 (2) | 1 (0) | 5 (5) | 1 (1) |
| Unknown | 317 (37) | 73 (62) | 93 (27) | 53 (33) | 5 (31) | 48 (34) |
| IHE does not offer student housing | 19 (3) | 7 (5) | 5 (2) | 2 (1) | 4 (3) | 1 (1) |
| 4. Made changes to campus common areas, n (%)‡ |  |  |  |  |  |  |
| Yes | 691 (78) | 72 (59) | 248 (83) | 127 (82) | 128 (94) | 116 (84) |
| Restricted room or area capacity | 508 (74) | 51 (74) | 187 (75) | 97 (76) | 99 (79) | 74 (63) |
| Use of physical guides (signage, floor stickers, etc.) | 428 (58) | 36 (50) | 150 (56) | 89 (70) | 78 (59) | 75 (65) |
| Use of physical barriers (furniture removal, partitions, etc.) | 397 (54) | 30 (44) | 139 (54) | 79 (61) | 76 (56) | 73 (64) |
| No | 1 (0) | 1 (1) | 0 (0) | 0 (0) | 0 (0) | 0 (0) |
| Unknown | 155 (22) | 45 (40) | 54 (17) | 25 (18) | 11 (6) | 20 (16) |
| 5. Made changes to campus dining halls, n (%)‡ |  |  |  |  |  |  |
| Yes | 642 (72) | 53 (44) | 234 (80) | 122 (80) | 119 (89) | 114 (84) |
| Increased grab and go options | 461 (73) | 40 (74) | 169 (73) | 87 (70) | 78 (68) | 87 (75) |
| Reduced capacity for indoor seating | 404 (67) | 32 (60) | 158 (70) | 77 (65) | 75 (71) | 62 (56) |
| Physical barriers or guides in lines and seating areas | 308 (45) | 24 (44) | 110 (42) | 56 (48) | 55 (47) | 63 (54) |
| Elimination of self service | 298 (48) | 28 (53) | 128 (55) | 47 (33) | 52 (47) | 43 (35) |
| No | 58 (3) | 8 (2) | 19 (2) | 14 (6) | 8 (5) | 9 (6) |
| Unknown | 147 (25) | 57 (54) | 49 (18) | 16 (14) | 12 (6) | 13 (10) |
| 6. Offered on-campus COVID-19 testing, n (%)‡ |  |  |  |  |  |  |
| Yes | 641 (67) | 48 (32) | 233 (73) | 125 (82) | 116 (85) | 129 (95) |
| Population eligible for COVID-19 testing |  |  |  |  |  |  |
| Residential students | 576 (90) | 39 (77) | 198 (90) | 112 (93) | 104 (92) | 123 (96) |
| All students | 543 (85) | 38 (74) | 181 (84) | 107 (90) | 100 (87) | 117 (91) |
| Staff | 400 (61) | 21 (45) | 12 (61) | 81 (61) | 79 (65) | 92 (72) |
| No minimum criteria to be tested | 399 (62) | 25 (39) | 141 (66) | 81 (68) | 55 (58) | 86 (67) |
| Scenarios when testing is required | 495 (83) | 43 (85) | 190 (88) | 94 (80) | 72 (70) | 96 (77) |
| Upon arrival for the semester | 363 (60) | 32 (62) | 155 (71) | 74 (54) | 44 (38) | 56 (43) |
| Random or scheduled surveillance testing of all students | 271 (47) | 21 (32) | 104 (51) | 52 (53) | 33 (34) | 61 (50) |
| Random or scheduled surveillance testing of residential students | 129 (20) | 2 (4) | 38 (21) | 27 (21) | 24 (20) | 38 (31) |
| Change in testing availability between academic terms |  |  |  |  |  |  |
| More available | 203 (32) | 19 (30) | 69 (32) | 43 (40) | 31 (27) | 41 (33) |
| Less available | 4 (0) | 0 (0) | 0 (0) | 3 (2) | 1 (1) | 0 (0) |
| No change | 106 (16) | 9 (23) | 36 (16) | 22 (15) | 21 (18) | 18 (13) |
| Unknown | 328 (51) | 20 (47) | 118 (51) | 57 (43) | 63 (54) | 70 (54) |
| No | 64 (9) | 15 (14) | 20 (7) | 13 (8) | 12 (9) | 4 (3) |
| Unknown | 142 (24) | 55 (54) | 59 (20) | 14 (10) | 11 (6) | 3 (2) |
| 7. Required masks or face coverings, n (%)‡ |  |  |  |  |  |  |
| Indoors | 784 (88) | 85 (68) | 288 (95) | 148 (96) | 133 (95) | 130 (96) |
| Students | 781 (88) | 83 (69) | 287 (95) | 148 (96) | 133 (95) | 130 (96) |
| Staff | 781 (88) | 84 (67) | 287 (94) | 148 (96) | 132 (94) | 130 (96) |
| Visitors | 750 (85) | 82 (65) | 275 (92) | 137 (91) | 128 (92) | 128 (95) |
| Outdoors | 723 (82) | 82 (65) | 262 (88) | 137 (88) | 120 (87) | 122 (89) |
| Students | 721 (82) | 81 (64) | 261 (87) | 137 (88) | 120 (87) | 122 (89) |
| Staff | 721 (82) | 82 (65) | 260 (87) | 137 (88) | 120 (87) | 122 (89) |
| Visitors | 694 (80) | 80 (63) | 252 (86) | 126 (83) | 117 (86) | 119 (87) |

\* NPIs derived from October 29, 2020 update of US Centers for Disease Control and Prevention Guidance for IHEs. Numbered items indicate each of the 7 broad NPIs being surveyed, followed by common modalities in which each NPI was implemented.
† Institution size category based on total students enrolled for credit as reported by the Integrated Postsecondary Education Data System.
‡ The universe of 1,181 private IHEs was stratified by institution size category. Based on sample size calculations using a 95% confidence interval with a 5% margin of error, 26% of eligible IHEs were randomly selected within each stratum (<1,000: 97 of 398 IHEs, 1,000 – 4,999: 161 of 625 IHEs, 5,000 – 9,999: 24 of 93 IHEs, 10,000 – 19,999: 12 of 47 IHEs, and ≥20,000: 6 of 18 IHEs). Weights represent the inverse of the sampling fractions. n (%) represents the unweighted number (n) and the weighted percent of the universe of 1,728 IHEs.

**Appendix Table 2.**
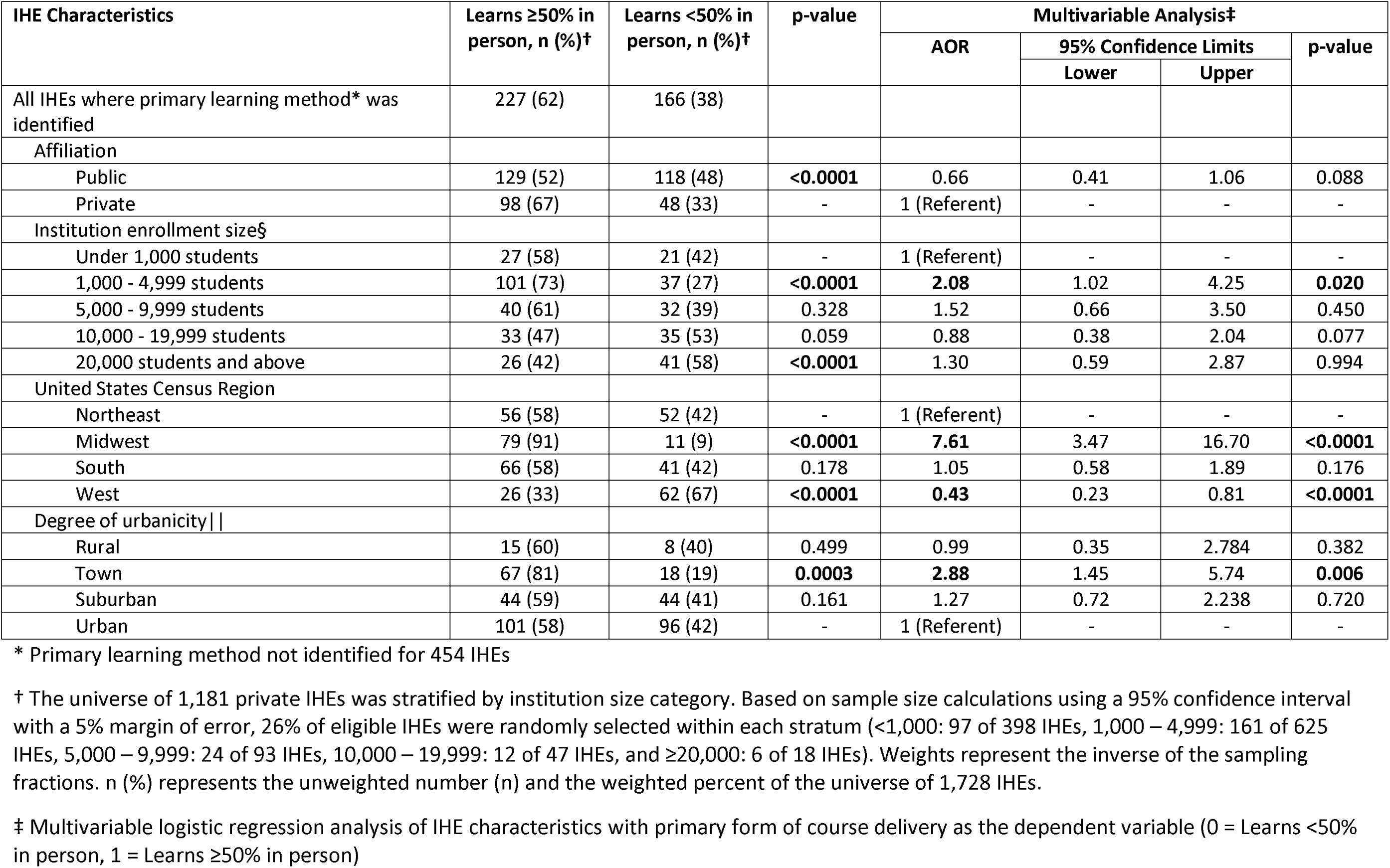
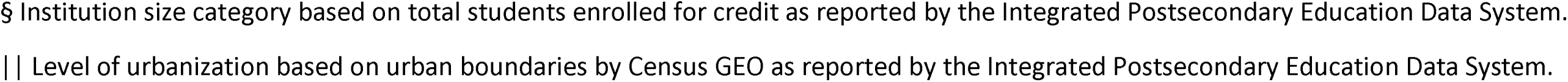
Characteristics of IHEs offering primarily in-person learning* (N=227) versus primarily distance learning (n=166), United States, Academic Year 2020–2021.

| IHE Characteristics | Learns ≥50% in person, n (%)† | Learns <50% in person, n (%)† | p-value | Multivariable Analysis‡ |  |  |  |
| --- | --- | --- | --- | --- | --- | --- | --- |
|  |  |  |  | AOR | 95% Confidence Limits |  | p-value |
|  |  |  |  |  | Lower | Upper |  |
| All IHEs where primary learning method* was identified | 227 (62) | 166 (38) |  |  |  |  |  |
| Affiliation |  |  |  |  |  |  |  |
| Public | 129 (52) | 118 (48) | **<0.0001** | 0.66 | 0.41 | 1.06 | 0.088 |
| Private | 98 (67) | 48 (33) | - | 1 (Referent) | - | - | - |
| Institution enrollment size§ |  |  |  |  |  |  |  |
| Under 1,000 students | 27 (58) | 21 (42) | - | 1 (Referent) | - | - | - |
| 1,000 - 4,999 students | 101 (73) | 37 (27) | **<0.0001** | **2.08** | 1.02 | 4.25 | **0.020** |
| 5,000 - 9,999 students | 40 (61) | 32 (39) | 0.328 | 1.52 | 0.66 | 3.50 | 0.450 |
| 10,000 - 19,999 students | 33 (47) | 35 (53) | 0.059 | 0.88 | 0.38 | 2.04 | 0.077 |
| 20,000 students and above | 26 (42) | 41 (58) | **<0.0001** | 1.30 | 0.59 | 2.87 | 0.994 |
| United States Census Region |  |  |  |  |  |  |  |
| Northeast | 56 (58) | 52 (42) | - | 1 (Referent) | - | - | - |
| Midwest | 79 (91) | 11 (9) | **<0.0001** | **7.61** | 3.47 | 16.70 | **<0.0001** |
| South | 66 (58) | 41 (42) | 0.178 | 1.05 | 0.58 | 1.89 | 0.176 |
| West | 26 (33) | 62 (67) | **<0.0001** | **0.43** | 0.23 | 0.81 | **<0.0001** |
| Degree of urbanicity\|\| |  |  |  |  |  |  |  |
| Rural | 15 (60) | 8 (40) | 0.499 | 0.99 | 0.35 | 2.784 | 0.382 |
| Town | 67 (81) | 18 (19) | **0.0003** | **2.88** | 1.45 | 5.74 | **0.006** |
| Suburban | 44 (59) | 44 (41) | 0.161 | 1.27 | 0.72 | 2.238 | 0.720 |
| Urban | 101 (58) | 96 (42) | - | 1 (Referent) | - | - | - |

\* Primary learning method not identified for 454 IHEs
† The universe of 1,181 private IHEs was stratified by institution size category. Based on sample size calculations using a 95% confidence interval with a 5% margin of error, 26% of eligible IHEs were randomly selected within each stratum (<1,000: 97 of 398 IHEs, 1,000 – 4,999: 161 of 625 IHEs, 5,000 – 9,999: 24 of 93 IHEs, 10,000 – 19,999: 12 of 47 IHEs, and ≥20,000: 6 of 18 IHEs). Weights represent the inverse of the sampling fractions. n (%) represents the unweighted number (n) and the weighted percent of the universe of 1,728 IHEs.
‡ Multivariable logistic regression analysis of IHE characteristics with primary form of course delivery as the dependent variable (0 = Learns <50% in person, 1 = Learns ≥50% in person)
§ Institution size category based on total students enrolled for credit as reported by the Integrated Postsecondary Education Data System.
|| Level of urbanization based on urban boundaries by Census GEO as reported by the Integrated Postsecondary Education Data System.

**Appendix Table 3.**
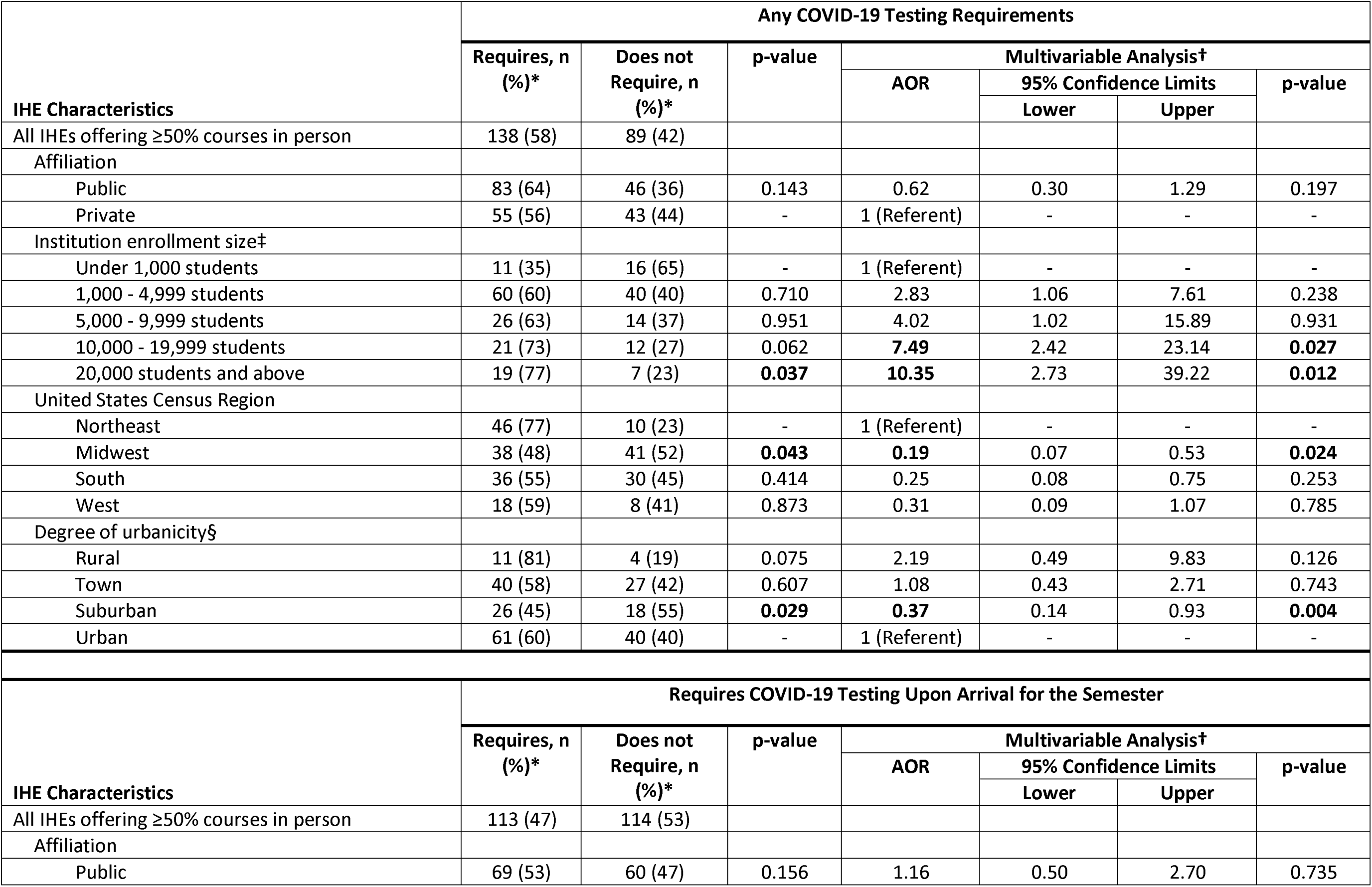
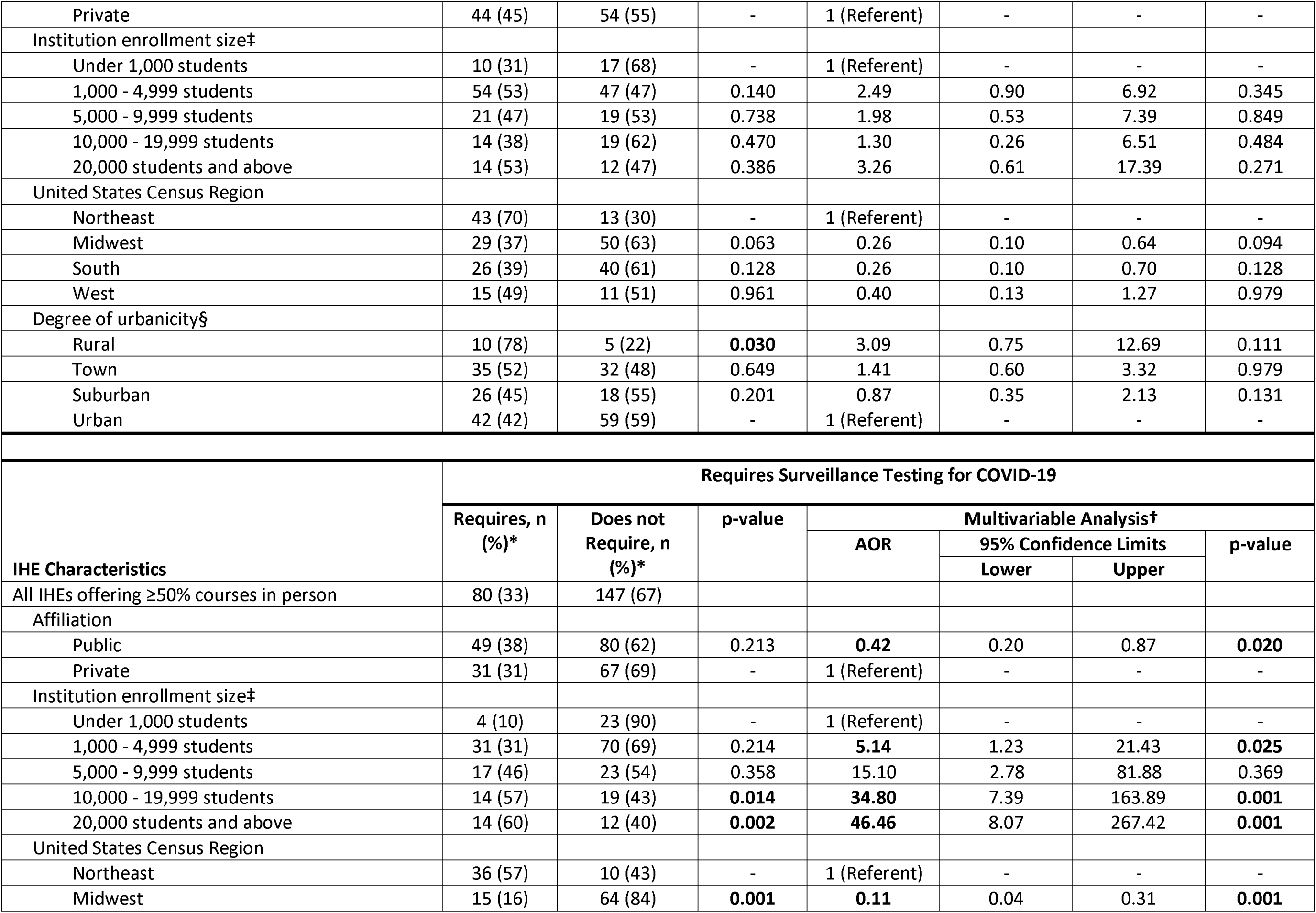
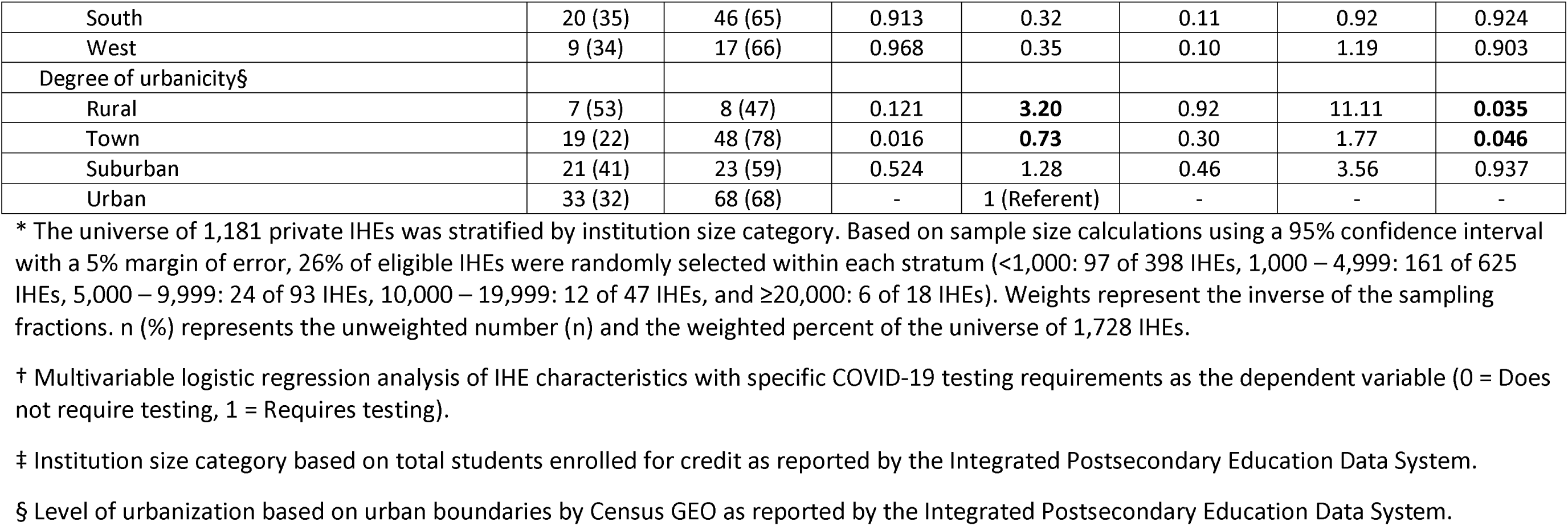
Characteristics of IHEs learning ≥50% in person (N=227) with specific COVID-19 testing requirements versus those without, United States, Academic Year 2020–2021.

**Appendix Figure 2.**
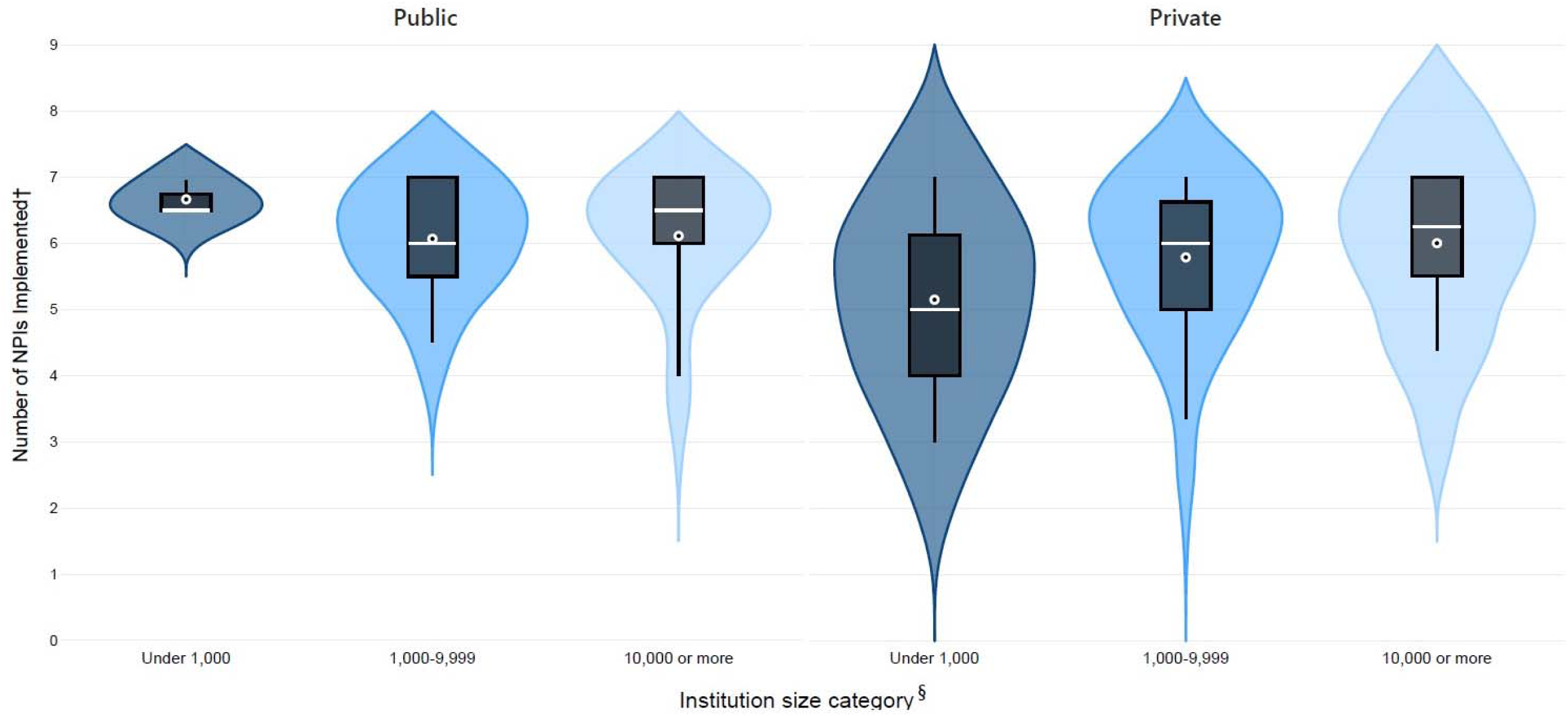
Distribution* of total number of nonpharmaceutical interventions (NPIs) implemented† by public (n=198) and private (n=98) institutions of higher education (IHEs) learning ≥50% in person‡ (N=227) within institution size categories^§^, United States, Academic Year 2020-2021. * Violin plot of total NPIs implemented kernel density distribution within institution size categories. Boxplot dot and line represent the mean and median number of NPIs implemented within each category, respectively. Generated in Microsoft Power BI. † NPIs derived from October 29, 2020 update of US Centers for Disease Control and Prevention Guidance for IHEs and summated ranging 0–7. ‡ 227 IHEs offer courses ≥50% in person. 166 IHEs offer courses <50% in person. Primary learning method not found for 454 IHEs. § Institution size category based on total students enrolled for credit as reported by the Integrated Postsecondary Education Data System.

## Acknowledgments

The authors would like to thank the data collection team, including in-house contractors (Esther Amoakohene, Tamara Cummings, Elisha Gilbert, Ferdous Jahan, and Zaneta Oliver) and ORISE fellows (Jeffrey Hodis and Olivia Shafer).

## References

1. Boehmer TK, DeVies J, Caruso E, et al. Changing Age Distribution of the COVID-19 Pandemic - United States, May-August 2020. MMWR Morbidity and mortality weekly report. 2020;69(39):1404–9. doi: 10.15585/mmwr.mm6939e1. PubMed PMID: 33001872.

2. Day T, Chang ICC, Chung CKL, et al. The Immediate Impact of COVID-19 on Postsecondary Teaching and Learning. Professional Geographer. 2021;73(1):1–13. doi: 10.1080/00330124.2020.1823864.

3. Guilamo-Ramos V, Benzekri A, Thimm-Kaiser M, et al. Reconsidering Assumptions of Adolescent and Young Adult Severe Acute Respiratory Syndrome Coronavirus 2 Transmission Dynamics. Clinical Infectious Diseases. 2020. doi: 10.1093/cid/ciaa1348.

4. Salvatore PP, Sula E, Coyle JP, et al. Recent Increase in COVID-19 Cases Reported Among Adults Aged 18-22 Years - United States, May 31-September 5, 2020. MMWR - Morbidity & Mortality Weekly Report. 2020;69(39):1419–24. doi: http://dx.doi.org/10.15585/mmwr.mm6939e4. PubMed PMID: 33006586.

5. Cipriano LE, Haddara WMR, Zaric GS, et al. Impact of university re-opening on total community COVID-19 burden. PLoS ONE [Electronic Resource]. 2021;16(8):e0255782. doi: https://dx.doi.org/10.1371/journal.pone.0255782. PubMed PMID: 34383796.

6. Cheng SY, Wang CJ, Shen AC, et al. How to Safely Reopen Colleges and Universities During COVID-19: Experiences From Taiwan. Ann Intern Med. 2020;173(8):638–41. doi: 10.7326/M20-2927. PubMed PMID: 32614638; PubMed Central PMCID: PMC7339040.

7. Benneyan JC, Gehrke C, Ilies I, et al. Potential Community and Campus Covid-19 Outcomes Under University and College Reopening Scenarios. MedRxiv : the Preprint Server for Health Sciences. 2020;02:02. doi: 10.1101/2020.08.29.20184366. PubMed PMID: 32908993.

8. Fox MD, Bailey DC, Seamon MD, et al. Response to a COVID-19 Outbreak on a University Campus - Indiana, August 2020. MMWR - Morbidity & Mortality Weekly Report. 2021;70(4):118–22. doi: 10.15585/mmwr.mm7004a3. PubMed PMID: 33507894; PubMed Central PMCID: PMC7842813.

9. Pollock BH, Kilpatrick AM, Eisenman DP, et al. Safe reopening of college campuses during COVID-19: The University of California experience in Fall 2020. PLOS ONE. 2021;16(11):e0258738. doi: 10.1371/journal.pone.0258738.

10. Guidance for institutions of higher education (IHEs) [Webpage]. Atlanta, GA: Centers for Disease Control and Prevention; 2022 [updated February 7, 2022]. Available from: https://www.cdc.gov/coronavirus/2019-ncov/community/colleges-universities/considerations.html.

11. Considerations for institutions of higher education [Webpage]. Atlanta, GA: Centers for Disease Control and Prevention; 2020 [updated October 29, 2020]. Available from: http://web.archive.org/web/20201029222917/https://www.cdc.gov/coronavirus/2019-ncov/community/colleges-universities/considerations.html.

12. The National Center for Education Statistics. Integrated Postsecondary Education Data System [Internet]. 2018-2019 [Accessed December 1, 2020]. Available from: https://nces.ed.gov/ipeds.

13. Qualls NL, Levitt AM, Kanade N, et al. Community mitigation guidelines to prevent pandemic influenza — United States, 2017. MMWR Recommendations and reports : Morbidity and mortality weekly report Recommendations and reports. 2017;66(1). https://stacks.cdc.gov/view/cdc/45220. PubMed PMID: cdc:45220.

14. 2010 Census Regions and Divisions of the United States [Internet]. US Census Bureau. 2010 [Accessed May 1, 2021]. Available from: https://www.census.gov/geographies/reference-maps/2010/geo/2010-census-regions-and-divisions-of-the-united-states.html.

15. COVID-19 Data Repository by the Center for Systems Science and Engineering (CSSE) [Internet]. Johns Hopkins University. 2020 [Accessed May 1, 2021]. Available from: https://github.com/CSSEGISandData/COVID-19.

16. Dong E DH, Gardner L. An interactive web-based dashboard to track COVID-19 in real time. Lancet Inf Dis. 20(5):533–4. doi: 10.1016/S1473-3099(20)30120-1.

17. Annual Resident Population Estimates for States and Counties: April 1, 2010 to July 1, 2019; April 1, 2020; and July 1, 2020 [Internet]. United States Census Bureau. 2020 [Accessed May 1, 2021]. Available from: https://www.census.gov/programs-surveys/popest/technical-documentation/research/evaluation-estimates/2020-evaluation-estimates/2010s-counties-total.html.

18. Ciol MA, Hoffman JM, Dudgeon BJ, et al. Understanding the use of weights in the analysis of data from multistage surveys. Arch Phys Med Rehabil. 2006;87(2):299–303. doi: 10.1016/j.apmr.2005.09.021. PubMed PMID: 16442990.

19. Iuliano AD, Reed C, Guh A, et al. Notes from the field: outbreak of 2009 pandemic influenza A (H1N1) virus at a large public university in Delaware, April-May 2009. Clin Infect Dis. 2009;49(12):1811–20. Epub 2009/11/17. doi: 10.1086/649555. PubMed PMID: 19911964.

20. Layde PM, Engelberg AL, Dobbs HI, et al. Outbreak of influenza A/USSR/77 at Marquette University. The Journal of infectious diseases. 1980;142(3):347–52. Epub 1980/09/01. doi: 10.1093/infdis/142.3.347. PubMed PMID: 7441004.

21. Cevasco KE, Roess AA, North HM, et al. Survival analysis of factors affecting the timing of COVID-19 non-pharmaceutical interventions by U.S. universities. BMC Public Health. 2021;21(1):1985. doi: 10.1186/s12889-021-12035-6.

22. Cevasco KE, North HM, Zeitoun SA, et al. COVID-19 observations and accompanying dataset of non-pharmaceutical interventions across U.S. universities, March 2020. PLOS ONE. 2020;15(10):e0240786. doi: 10.1371/journal.pone.0240786.

23. Wilson E, Donovan CV, Campbell M, et al. Multiple COVID-19 Clusters on a University Campus - North Carolina, August 2020. MMWR - Morbidity & Mortality Weekly Report. 2020;69(39):1416–8. doi: 10.15585/mmwr.mm6939e3. PubMed PMID: 33001871; PubMed Central PMCID: PMC7537562.

24. MacIntyre CR, Costantino V, Bian L, et al. Effectiveness of facemasks for opening a university campus in Mississippi, United States – a modelling study. Journal of American College Health. 2021:1–6. doi: 10.1080/07448481.2020.1866579.

25. Barrios LC, Riggs MA, Green RF, et al. Observed face mask use at six universities — United States, September–November 2020. 2021;70(6):208–11. doi: 10.15585/mmwr.mm7006e1. PubMed PMID: 33571175; PubMed Central PMCID: PMC7877579.

26. Chang T, Draper JM, Van den Bout A, et al. A method for campus-wide SARS-CoV-2 surveillance at a large public university. PLOS ONE. 2021;16(12):e0261230. doi: 10.1371/journal.pone.0261230.

27. Schultes O, Clarke V, Paltiel AD, et al. COVID-19 Testing and Case Rates and Social Contact Among Residential College Students in Connecticut During the 2020-2021 Academic Year. JAMA Netw Open. 2021;4(12):e2140602-e. doi: 10.1001/jamanetworkopen.2021.40602.

28. Leidner AJ, Barry V, Bowen VB, et al. Opening of Large Institutions of Higher Education and County-Level COVID-19 Incidence - United States, July 6-September 17, 2020. MMWR - Morbidity & Mortality Weekly Report. 2021;70(1):14–9. doi: 10.15585/mmwr.mm7001a4. PubMed PMID: 33411699; PubMed Central PMCID: PMC7790156.

29. Lehnig CL, Oren E, Vaidya NK. Effectiveness of alternative semester break schedules on reducing COVID-19 incidence on college campuses. Scientific Reports. 2022;12(1):2116. doi: 10.1038/s41598-022-06260-1.

30. Best A, Singh P, Ward C, et al. The impact of varying class sizes on epidemic spread in a university population. R. 2021;8(6):210712. doi: https://dx.doi.org/10.1098/rsos.210712. PubMed PMID: 34150319.

31. Connor C. Factors in the probability of covid-19 transmission in university classrooms. Numeracy. 2020;13(2):1–15. doi: 10.5038/1936-4660.13.2.1368.

32. Ambatipudi M, Carrillo Gonzalez P, Tasnim K, et al. Risk quantification for SARS-CoV-2 infection through airborne transmission in university settings. J Occup Environ Hyg. 2021:1–19. doi: https://dx.doi.org/10.1080/15459624.2021.1985725. PubMed PMID: 34569919.

33. Ascione F, De Masi RF, Mastellone M, et al. The design of safe classrooms of educational buildings for facing contagions and transmission of diseases: A novel approach combining audits, calibrated energy models, building performance (BPS) and computational fluid dynamic (CFD) simulations. Energy & Buildings. 2021;230:110533. doi: 10.1016/j.enbuild.2020.110533. PubMed PMID: 33052169; PubMed Central PMCID: PMC7543903.

34. Murray AT. Planning for classroom physical distancing to minimize the threat of COVID-19 disease spread. PLoS ONE [Electronic Resource]. 2020;15(12):e0243345. doi: https://dx.doi.org/10.1371/journal.pone.0243345. PubMed PMID: 33275618.

35. Education SCoH. The Impact of COVID-19 on Higher Education. MAPS Project: The University of Utah, 2021.

36. Katz VS, Jordan AB, Ognyanova K. Digital inequality, faculty communication, and remote learning experiences during the COVID-19 pandemic: A survey of U.S. undergraduates. PLoS ONE [Electronic Resource]. 2021;16(2):e0246641. doi: 10.1371/journal.pone.0246641. PubMed PMID: 33566832; PubMed Central PMCID: PMC7875367.

37. Unger S, Meiran WR. Student Attitudes towards Online Education during the COVID-19 Viral Outbreak of 2020: Distance Learning in a Time of Social Distance. International Journal of Technology in Education and Science. 2020;4(4):256–66. doi: 10.46328/ijtes.v4i4.107. PubMed PMID: 2488216723; EJ1271377.

38. Mostafa S, Cousins-Cooper K, Tankersley B, et al. The impact of COVID-19 induced emergency remote instruction on students’ academic performance at an HBCU. PLOS ONE. 2022;17(3):e0264947. doi: 10.1371/journal.pone.0264947.

39. Benjamin Collins JHF, Cassandria Dortch. The COVID-19 Pandemic and Institutions of Higher Education: Contemporary Issues. Congressional Research Service, 2021 Contract No.: R46666.

40. Losina E, Leifer V, Millham L, et al. College Campuses and COVID-19 Mitigation: Clinical and Economic Value. Ann Intern Med. 2021;174(4):472–83. Epub 2020/12/21. doi: 10.7326/M20-6558. PubMed PMID: 33347322.

41. Adams GB, Shannon J, Shannon S. “Return to University Campuses Associated with 9% Increase in New COVID-19 Case Rate”. medRxiv. 2020. doi: https://doi.org/10.1101/2020.10.13.20212183.

42. Doyle K, Teran RA, Reefhuis J, et al. Multiple Variants of SARS-CoV-2 in a University Outbreak After Spring Break - Chicago, Illinois, March-May 2021. MMWR - Morbidity & Mortality Weekly Report. 2021;70(35):1195–200. doi: https://dx.doi.org/10.15585/mmwr.mm7035a3. PubMed PMID: 34473687.

43. Moreno G, Braun K, Pray I, et al. Characterizing SARS-CoV-2 spread on college campuses. Journal of the International AIDS Society. 2021;24(SUPPL 1):107–8. doi: http://dx.doi.org/10.1002/jia2.25659. PubMed PMID: 634543317.

44. Vang KE, Krow-Lucal ER, James AE, et al. Participation in Fraternity and Sorority Activities and the Spread of COVID-19 Among Residential University Communities - Arkansas, August 21-September 5, 2020. MMWR - Morbidity & Mortality Weekly Report. 2021;70(1):20–3. doi: http://dx.doi.org/10.15585/mmwr.mm7001a5. PubMed PMID: 33411698.

45. AAMC Recommendations for COVID-19 Testing: The Current State and The Way Forward. The Way Forward on COVID-19 [Internet]. 2020 2/10/2022. Available from: https://www.aamc.org/covidroadmap/testing.

46. Paltiel AD, Zheng A, Walensky RP. Assessment of SARS-CoV-2 Screening Strategies to Permit the Safe Reopening of College Campuses in the United States. JAMA Netw Open. 2020;3(7):e2016818-e. doi: 10.1001/jamanetworkopen.2020.16818. PubMed PMID: 32735339.

47. Oran DP, Topol EJ. Prevalence of Asymptomatic SARS-CoV-2 Infection : A Narrative Review. Ann Intern Med. 2020;173(5):362–7. Epub 2020/06/03. doi: 10.7326/M20-3012. PubMed PMID: 32491919.

48. Fung IC, Cheung CN, Handel A. SARS-CoV-2 Viral and Serological Testing When College Campuses Reopen: Some Practical Considerations. Disaster Medicine & Public Health Preparedness. 2020:1–5. doi: 10.1017/dmp.2020.266. PubMed PMID: 32713384; PubMed Central PMCID: PMC7450242.

49. Goyal R, Hotchkiss J, Schooley RT, et al. Evaluation of SARS-CoV-2 transmission mitigation strategies on a university campus using an agent-based network model. Clinical Infectious Diseases. 2021;19:19. doi: 10.1093/cid/ciab037. PubMed PMID: 33462589; PubMed Central PMCID: PMC7929036.

